# Clinical Ordering Practices of the SARS-CoV-2 Antibody Test at a Large Academic Medical Center

**DOI:** 10.1101/2020.07.12.20152165

**Authors:** Joesph R. Wiencek, Carter L. Head, Costi D. Sifri, Andrew S. Parsons

## Abstract

**Background:** The novel severe acute respiratory coronavirus 2 (SARS-CoV-2) that causes COVID-19 originated in December 2019 and has now infected over 3 million people in the United States. In Spring of 2020, private laboratories and some hospitals began antibody testing despite lacking evidence-based guidance.

**Objective:** To describe clinician-described indications for SARS-CoV-2 antibody testing, including cost implications, immediately following testing availability.

**Design:** Retrospective chart review of patients who received antibody testing from May 14, 2020 to June 15, 2020.

**Setting:** A large academic medical center, one of the first in the US to provide antibody testing capability to individual clinicians.

**Patients:** 447 consecutive patients who received SARS-CoV-2 antibody testing.

**Measurements:** Clinician-described indications for SARS-CoV-2 antibody testing, cost implications, and comparison with current expert-based guidance from the IDSA and CDC.

**Results:** Of 444 individual antibody test results meeting inclusion criteria, the two most commonly described indications for ordering the antibody test, apart from public health epidemiology studies (n=223), were for patients with a now resolved COVID-19 compatible illness (n=105) with no previous molecular testing and in asymptomatic patients believed to have had a past exposure or contact with a person with COVID-19 compatible illness (n=60). The rate of positive SARS-CoV-2 antibody testing among those indications consistent with current IDSA and CDC guidance was 17% compared with 5% (p<0.0001) among those indications inconsistent with current IDSA and CDC guidance. Total cost estimates ranged from $57,720 to $97,680, of which 42% was for testing inconsistent with current expert-based guidance.

**Limitations:** The duration of antibody response following infection is unclear and asymptomatic individuals may not develop a positive antibody response.

**Conclusions:** Our findings demonstrate a dissociation between clinician described indications for testing and expert-based guidance and a significantly different rate of positive testing between these two groups. Clinical curiosity and patient preference appear to have played a significant role in testing decisions and substantially contributed to testing costs.

## Introduction

Coronavirus disease 19 (COVID-19) is an international crisis that continues to influence every facet of human life. The novel severe acute respiratory coronavirus 2 (SARS-CoV-2) that causes COVID-19 manifested itself in December 2019.(1),(2) As of June 27, 2020, over 3 million people in the United States (US) and over 11 million people worldwide have been confirmed with the SARS-CoV-2 infection.(3) Over the past few months, the key to detection as well as understanding the spread of the virus has been primarily through evolving testing strategies.(4)

There are two main clinical laboratory tests being used to monitor the widespread infection of the SARS-CoV-2 virus.(4) The first diagnostic tests for COVID-19 were based on the detection of viral genetic material for SARS-CoV-2. These molecular diagnostic tests (i.e., reverse transcription polymerase chain reaction, RT-PCR) remain the gold standard for diagnosing active viral infection in symptomatic COVID-19 cases.(5) Several weeks after the molecular tests were implemented, antibody tests to the SARS-CoV-2 virus became available. Over 200 antibody tests (e.g., IgG, IgM and IgG/IgM combo) flooded the market and were distributed without being properly vetted through the typical Food and Drug Administration (FDA) process.(6) This led to general concerns about various antibody test accuracy, prompting the FDA to modify its stance about data review and to now require all in vitro diagnostic test manufacturers to submit their tests through the FDA Emergency Use Authorization pathway.(7) The utility and limitations of the antibody test have been at the forefront of discussions in the medical and lay communities.(8,9),(10)

The clinical and public health necessity for ordering the SARS-CoV-2 antibody test remains poorly described. To this point, the test has primarily been used in epidemiological studies to determine disease burden.(11,12) For instance, a recent analysis of SARS-CoV-2 antibody test results from nearly 12,000 serum samples collected as part of routine or sick care (but unrelated to COVID-19) suggested that the COVID-19 burden may be more than ten times greater than previously thought.(12) Yet, the antibody response in infected patients remains largely unknown, and the clinical value of antibody testing has not been fully demonstrated.(13,14) Given the current lack of evidence-based guidance, guidelines from the Infectious Diseases Society of America (IDSA) based on expert opinion focused on four situations where the detection of antibodies could be helpful such as public health surveillance and identification of convalescent plasma donors.(15) Similarly, the Centers for Disease Control and Prevention (CDC) released interim guidance that urged healthcare professionals to not use antibody tests as a means to diagnosis a patient with COVID-19.(16) Despite lacking evidence-based guidance, private laboratories and some hospitals began antibody testing in early spring of 2020.(17)

Widespread availability of the SARS-CoV-2 antibody test with limited guidance on appropriate testing indications within the clinical environment may lead to significant healthcare waste as the pandemic continues. Excessive laboratory testing is a known driver of healthcare waste and can negatively impact clinical outcomes.(18) The SARS-CoV-2 antibody test is currently estimated to cost between $30 and $50, a price that increases to between $120 and $175 dollars when administration costs are included.(19) America’s Health Insurance Plans (AHIP) estimates that widespread antibody testing could cost the US between $5 and $19 billion.(19) Yet, incomplete evidence on how the test currently is or should be used limits an accurate estimate of cost in a time when the resource implications of widespread antibody testing are of upmost importance.

Clinicians and insurance providers are seeking answers as to the value of SARS-CoV-2 antibody testing. A number of US hospitals have issued perspective communications hoping to address employee and patient concerns related to a lack of guidance on the necessity of antibody testing.(20) Similarly, infectious disease and public health leaders recently published an opinion piece expressing concerns over the lack of specific clinical indications following the establishment of SARS-CoV-2 antibody testing in England.(21) In a recent letter to Secretary of Health and Human Services Alex Azar, the National Association of Insurance Commissioners (NAIC) wrote, “Centers for Medicare and Medicaid Services (CMS) guidance has indicated testing should be covered by insurers when medically necessary, but there are still many questions about the application of medical necessity standards and FDA guidance is less clear about the value of various tests. States, local governments, employers, and carriers would benefit from a consistent and clear message about what tests are approved for what uses and in what circumstances they should be covered by insurers.”(22) This uncertainty around coverage could undermine Congress’ testing mandate and may lead to significant costs for hospitals and private laboratories that could ultimately be passed on to patients.

Herein, we describe the indications for SARS-CoV-2 antibody testing immediately following testing availability at a large academic medical center. As one of the first medical centers in the US to provide antibody testing capability to individual clinicians, we hope our experience serves as a guide for clinicians at other medical centers utilizing SARS-CoV-2 antibody testing and informs policy-makers aiming to define testing indications that promote high-value care to patients and the public at large.

## Methods

### Data Collection

The University of Virginia Health System (UVAHS) began SARS-CoV-2 antibody testing on May 14, 2020. Antibody testing was performed in our central clinical laboratory on the Abbott Architect i2000 analyzer utilizing the EUA SARS-CoV-2 IgG antibody immunoassay. Before any patients were tested, the SARS-CoV-2 IgG antibody test was validated similarly as previously described.^20^ We conducted a retrospective chart review of patients who received antibody testing at UVAHS from May 14, 2020 to June 15, 2020 to identify the indications for testing as described by the ordering clinician in the electronic medical record (EMR). Chart review was performed under a protocol that was approved by the University of Virginia Institutional Review Board (IRB-HSR #13310).

To identify patients to be included in the retrospective chart review, we developed an automated report that identified SARS-CoV-2 antibody test orders within the EMR. This report included fields for order identification number, date of order, ordering clinician, patient location, laboratory procedure number, medical record number (MRN), test result, result time, and result status. Prisoners, patients tested for test validation, and patients with charts that were locked from outside review due to privacy concerns were excluded.

When available, the patient MRN was used to access and review patient charts within the EMR to identify the indication used by the clinician to order the SARS-CoV-2 antibody test. To do this, we reviewed recent patient encounter documentation including patient charts, MyChart communications, and laboratory orders. We looked for documentation by the ordering clinician indicating the purpose for ordering the SARS-CoV-2 antibody test reviewing as far back as January 1, 2020. If there was not enough information available for the reviewer to identify the indication the ordering clinician was using, then a ‘no indication provided’ label was assigned to that test.

Beyond the indication used for antibody testing, we collected additional information including age, sex, timing for onset of symptoms as reported by the patient, as well as any records of COVID-19 viral RT-PCR tests ordered prior to the SARS-CoV-2 antibody test with the associated date and result of the test. Due to the retrospective focus of the study, we did not include RT-PCR tests in our data that occurred on dates after the SARS-CoV-2 antibody test.

A subset of the laboratory orders included in the automated report did not have an associated MRN, preventing us from performing chart reviews to identify the indication for antibody testing. However, using the ordering clinician information included in the report, we were able to identify the indication for testing without reviewing the chart. Within our study, there were two scenarios that required this approach. The first was a small subset of SARS-CoV-2 tests ordered for efficacy evaluation of this newly developed antibody test. These orders were excluded from our data set (n=3). The second scenario was related to epidemiologic studies evaluating the prevalence of COVID-19 by public health entities (n=222). This second group of tests was included in our analysis as epidemiologic studies fall within the current guidelines for ordering antibody testing.

### Statistical Analysis

All data was de-identified and collected into Microsoft Excel. The range, mean, standard deviation, and median were determined. The unpaired t test, assuming equal variance with an alpha value of 0.05, was used to compare means of two unmatched groups, those testing indications consistent with IDSA or CDC guidance and those testing indications inconsistent with such guidance.

### Review of Evidence

Current IDSA and CDC expert-based recommendations for antibody testing were reviewed to formulate a list of possible testing indications (Table 1). Indications noted in the EMR that were not included in the CDC or IDSA guidelines were added to the list (Table 1). There were 11 indications in total. Indications A, E, H, and K were provided by the IDSA and indications A, E, H, and F were provided by the CDC. Recommendations B, C, D, G, I, J were based on chart review. Once a clinician-described indication was identified through chart review, the indication was matched to the list of testing indications developed by our team. The positive antibody result rate was then calculated for these patients.

**Table 1.**
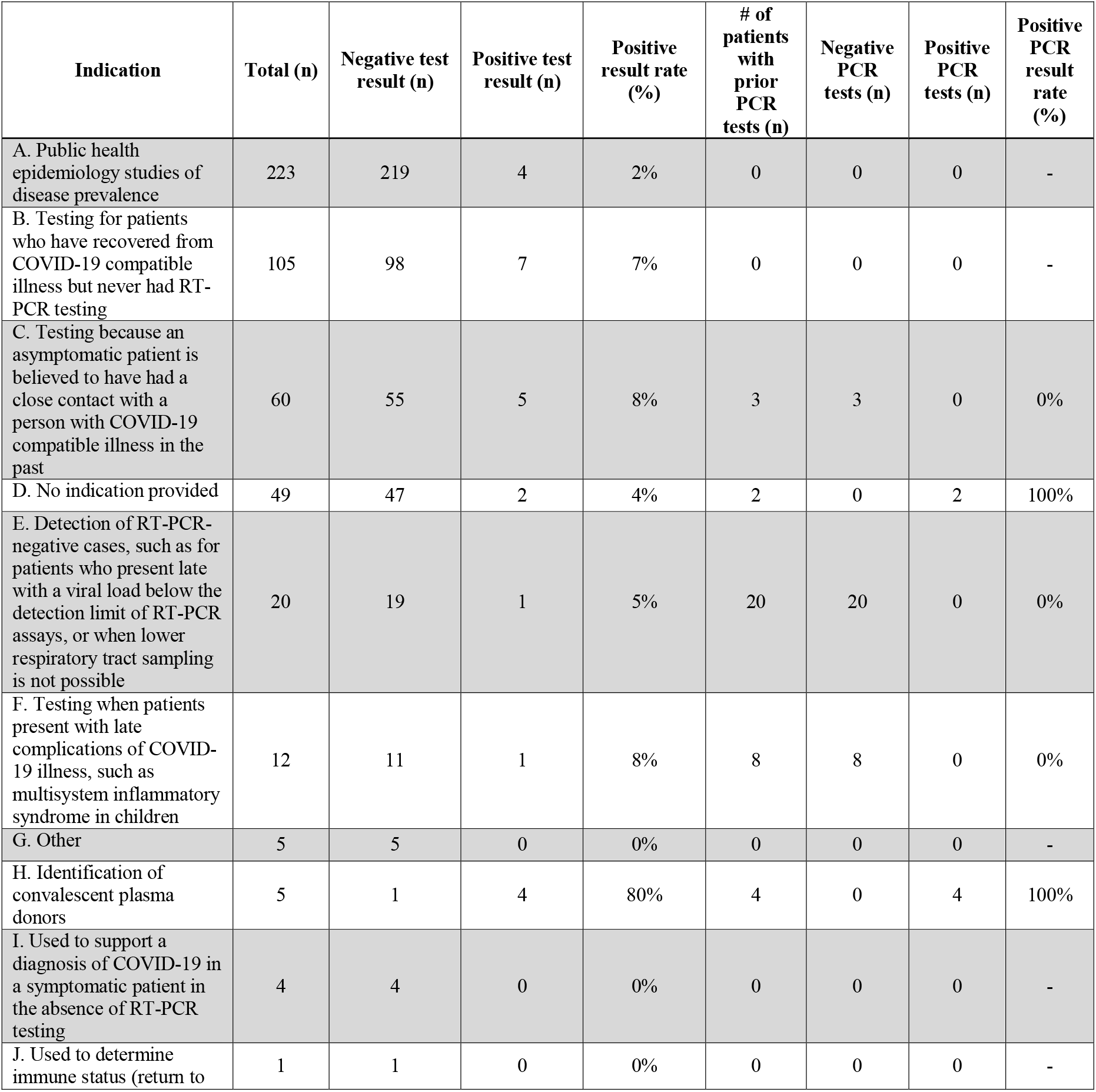
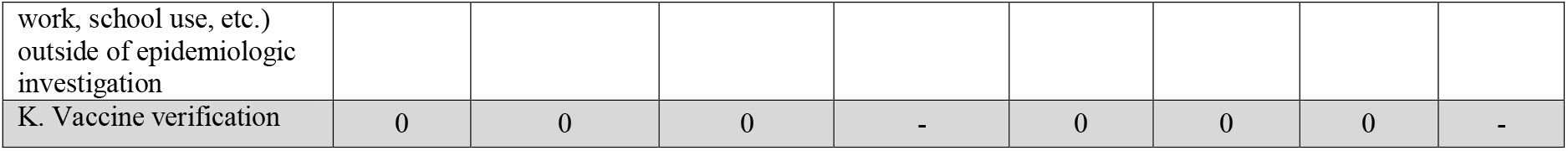
SARS-CoV-2 antibody test and SARS-CoV-2 RT-PCR test results by indication used to order the antibody test. The indications used to order SARS-CoV-2 antibody tests are listed in column 1. The total number of tests using each indication are listed in column 2. In columns 3 and 4, the number of negative and positive antibody test results for each testing indication are listed, and the positive antibody result rate as a percentage is in column 5. Column 6 lists the number of patients who had viral RT-PCR testing before antibody testing. The count of negative RT-PCR results, positive RT-PCR results, and positive result rate as a percentage of RT-PCR testing are listed in columns 7, 8, and 9 respectively.

### Cost Analysis

An estimated cost analysis of the SARS-CoV-2 antibody test was performed utilizing all reviewed tests between May 14, 2020 to June 15, 2020. Test and test administration (i.e., office visit) cost data from the AHIP were used to determine total costs in both low and high cost situations (Table 2).(19) This information was further used to determine the breakdown costs associated with antibody testing within or outside current expert-based guidance.

**Table 2.** High and low-cost scenarios for different segments of SARS-CoV-2 antibody testing. In Table 2, cost estimates for SARS-CoV-2 antibody testing are outlined. Different testing segments were identified based on the indications used to order the antibody test. The segments included: all testing, testing outside expert-based guidance, testing within expert-based guidance, testing within expert-based guidance excluding public health epidemiology studies, and testing for public health epidemiology studies. The total tests for each segment have been listed in line 2. Next, low and high testing costs were identified. This cost included the antibody test cost and cost to administer the test (i.e. office visit). A low and high cost estimate for each item was used. Our low-end estimate utilizes the AHIP’s low-end Medicare testing estimates, and our high-end estimate utilizes AHIP’s high-end commercial testing estimates.(19) Once low and high total costs to perform a single test were identified, total low and high testing costs for each segment were calculated by multiplying the total cost for a single test by the total number of tests in a segment (line 6). Next, the positive result rate of each segment was calculated as the number of positive test results out of total tests for a segment. This is listed as a percentage in line 7. Finally, the cost per positive test for low and high cost scenarios of each segment were calculated (line 8). This was done by dividing the total cost of testing by the number of positive test results in each segment.

| Test segment | All reviewed tests performed<br>from 5/14/20 to 6/15/20 |  | All reviewed tests performed<br>from 5/14/20 to 6/15/20<br>outside current guidelines |  | All reviewed tests performed<br>from 5/14/20 to 6/15/20 within<br>current guidelines |  |
| --- | --- | --- | --- | --- | --- | --- |
| **Total tests** | 444 | 444 | 186 | 186 | 258 | 258 |
| **Cost scenario** | Low Cost | High Cost | Low Cost | High Cost | Low Cost | High Cost |
| **Antibody test**<br>**cost (US\$)** | \$ 40.00 | \$ 65.00 | \$ 40.00 | \$ 65.00 | \$ 40.00 | \$ 65.00 |
| **Test**<br>**administration**<br>**cost (US\$)** | \$ 90.00 | \$ 155.00 | \$ 90.00 | \$ 155.00 | \$ 90.00 | \$ 155.00 |
| **Total cost**<br>**(US\$)** | \$ 57,720.00 | \$ 97,680.00 | \$ 24,180.00 | \$ 40,920.00 | \$ 33,540.00 | \$ 56,760.00 |
| **Positive test**<br>**rate (%)** | 5% | 5% | 5% | 5% | 4% | 4% |
| **Cost per**<br>**positive test**<br>**(US\$)** | \$ 2,886.00 | \$ 4,884.00 | \$ 2,418.00 | \$ 4,092.00 | \$ 3,354.00 | \$ 5,676.00 |

**Table 2 Legend.** In Table 2, cost estimates for SARS-CoV-2 antibody testing are outlined.
| Test segment | All reviewed tests performed from 5/14/20 to 6/15/20 for public health epidemiology studies* |  | All reviewed tests performed from 5/14/20 to 6/15/20 within guidelines excluding public health epidemiology studies |  |
| --- | --- | --- | --- | --- |
| Total tests | 223 | 223 | 35 | 35 |
| Cost scenario | Low Cost | High Cost | Low Cost | High Cost |
| Antibody test cost (US\$) | \$ 40.00 | \$ 65.00 | \$ 40.00 | \$ 65.00 |
| Test administration cost (US\$) | \$ 90.00 | \$ 155.00 | \$ 90.00 | \$ 155.00 |
| Total cost (US\$) | \$ 28,990.00 | \$ 49,060.00 | \$ 4,550.00 | \$ 7,700.00 |
| Positive test rate (%) | 2% | 2% | 17% | 17% |
| Cost per positive test (US\$) | \$ 7,247.50 | \$ 12,265.00 | \$ 758.33 | \$ 1,283.33 |

\*Testing costs may vary within public health epidemiology studies

## Results

We reviewed a total of 449 SARS-CoV-2 antibody tests collected for 447 patients over a one month time period. Five patients were excluded. Half of the remaining patients (n=220) were stratified by age and sex. Males (n=93) and females (n=127) were further separated into five distinct age groups, (0-20, 21-40 41-60, 61-80, and >80 years). The age brackets from 41-60 and 61-80 contained the most antibody tests for females (n=48) and males (n=35), respectively. Race and ethnicity information were not collected for this study as these variables were often inconsistent or missing in the EMR. The remaining (n=222) patients were associated with public health epidemiology studies and the EMR did not contain any demographic or additional patient information.

All patients were separated into distinct testing indications. This allowed nine separate categories for ordering indications for the SARS-CoV-2 antibody test and two additional groupings broken down as “other” or “no indication provided” (Figure 1). Each ordering indication was further classified by antibody test result and positive result rate as well as molecular test information, when available (Table 1). The two most commonly described indications for ordering the antibody test were epidemiologic studies (n=223, 50%) and patients who have resolved COVID-19 compatible illness (n=105, 24%) but never had a molecular test. The highest percentage positive result rate (80%) was to identify convalescent plasma donors. No testing was done on patients to determine vaccine response (i.e., indication K), owing to the fact that vaccines to the SARS-CoV-2 virus are not currently available. Overall, the rate of positive SARS-CoV-2 antibody testing among all indications ranged from 0 to 80% with a median of 5%. Notably, when testing for epidemiological studies was removed, the rate of positive SARS-CoV-2 antibody testing among those indications consistent with current IDSA and CDC guidance was 17%. In comparison, the rate of positive SARS-CoV-2 antibody testing among those indications inconsistent with current IDSA and CDC guidance was 5% (p<0.0001).

**Figure 1.**
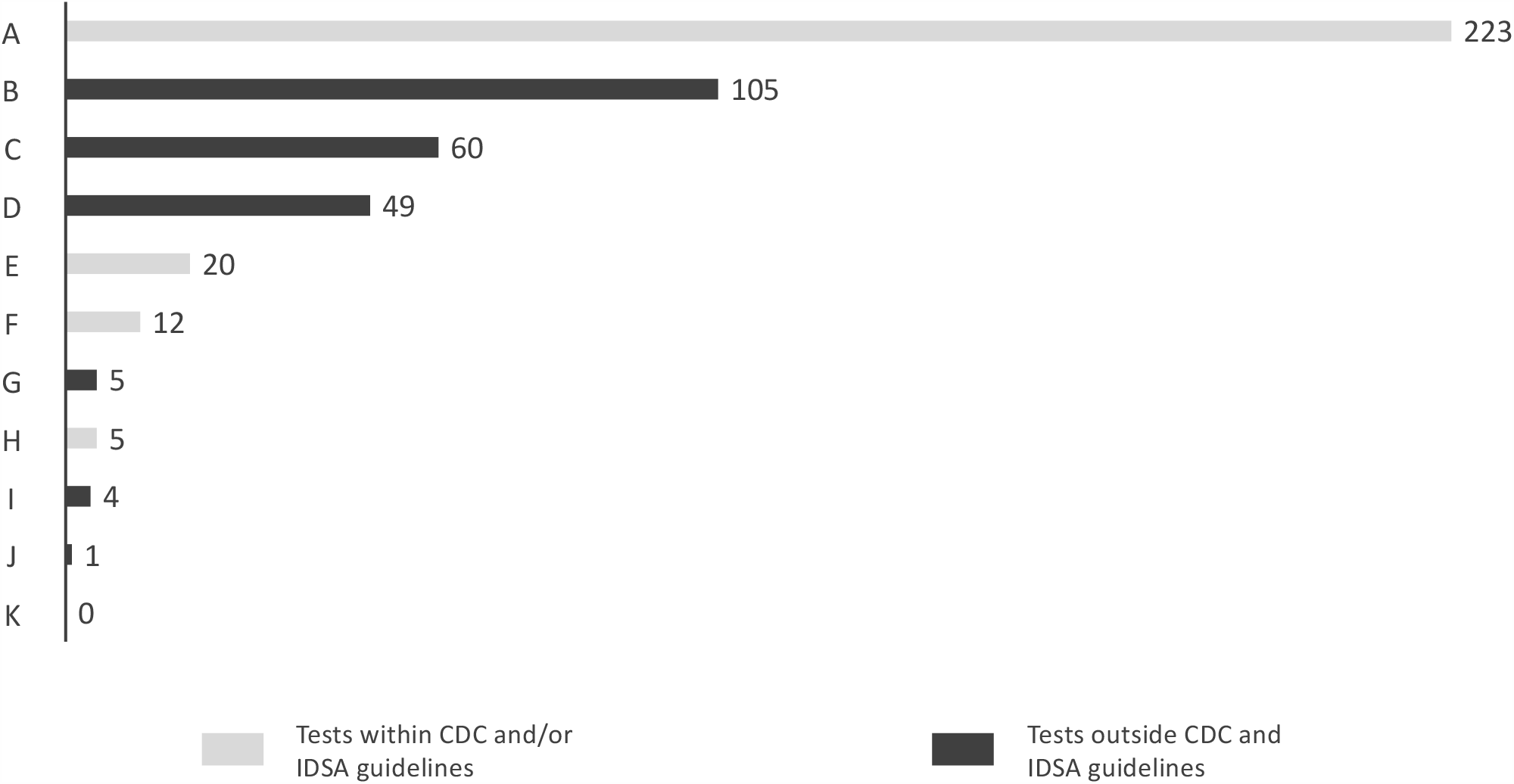
Incidence of SARS-CoV-2 antibody tests by indication used to order the test. The y axis reflects the indication used to order SARS-CoV-2 antibody test (A-K) and the x-axis represents the number of tests. Indications were defined as **A:** Public health epidemiology studies of disease prevalence. **B:** Testing for patients who have recovered from COVID-19 compatible illness but never had RT-PCR testing. **C:** Testing because an asymptomatic patient is believed to have had a close contact with a person with COVID-19 compatible illness in the past. **D:** No indication provided. **E:** Detection of RT-PCR-negative cases, such as for patients who present late with a viral load below the detection limit of RT-PCR assays, or when lower respiratory tract sampling is not possible. **F:** Testing when patients present with late complications of COVID-19 illness, such as multisystem inflammatory syndrome in children. **G:** Other indication used. **H:** Identification of convalescent plasma donors. **I:** Used to support a diagnosis of COVID-19 in a symptomatic patient in the absence of RT-PCR testing. **J:** Used to determine immune status (return to work, school use, etc.) outside of epidemiologic investigation. **K:** Vaccine verification.

Time periods were identified by chart review for patients who self-reported COVID-19 like symptoms. Sixty-six percent of patients (n=146) reported having symptoms either in a specific month as far back as November 2019 or indicated more broadly that they felt ill in late 2019 or early 2020 (Figure 2). From January 2020 to May 2020, the positive result rate ranged from 7% to 13% with a median of 12%. Positive result rates were zero in all other reported time frames.

**Figure 2.**
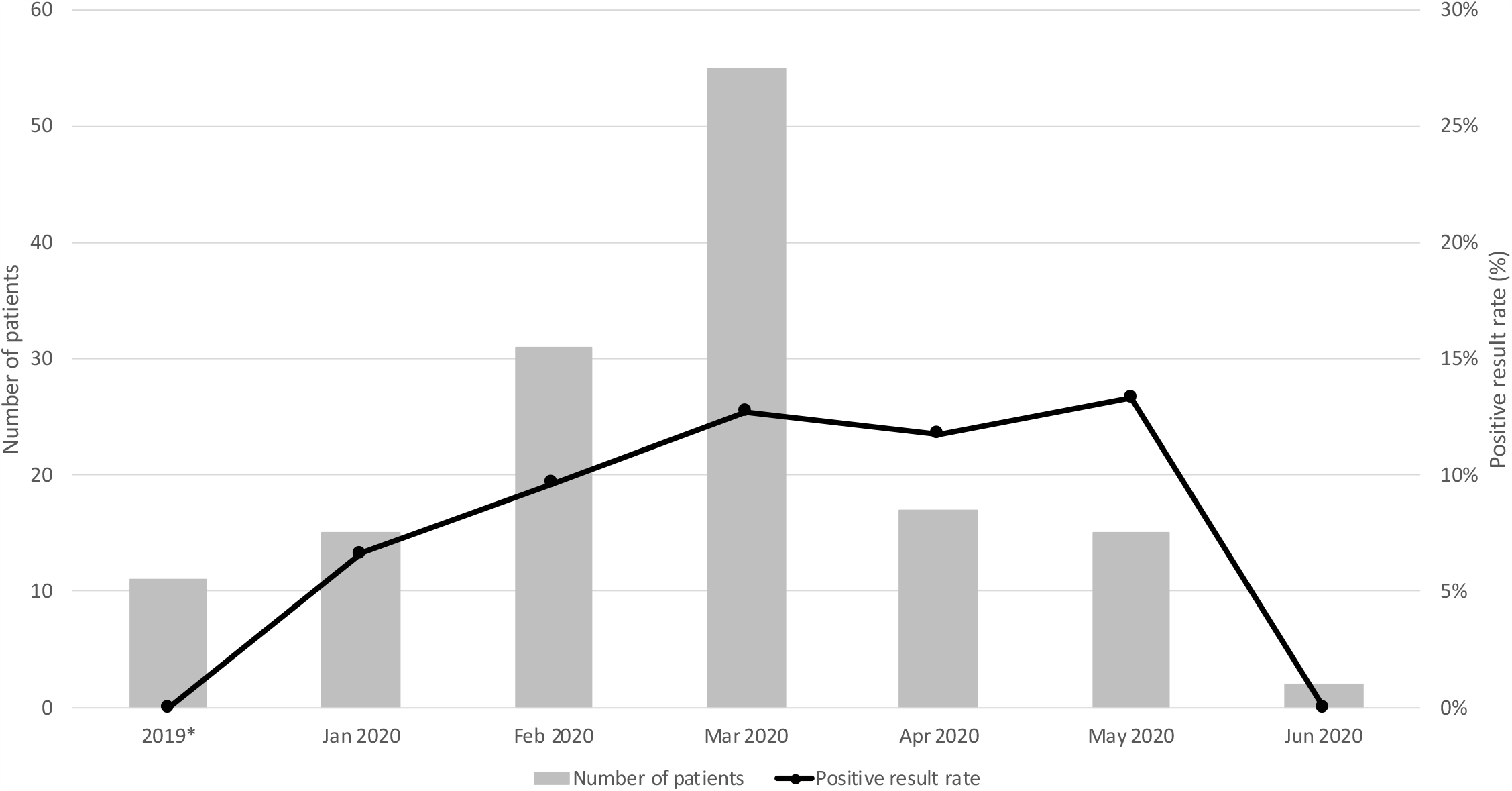
Patient reported frequency of onset COVID-19 like symptoms and positive result rate. The left and right y-axis represents the number of patients who reported COVID-19 like symptoms and positive result rate as a percentage, respectively. The x-axis includes the month and year when patients reported COVID-19 like symptoms. *Includes patients reporting symptoms at any time within the year 2019

Total cost estimates for all tests performed during the first month of testing ranged from $57,720 to $97,680 (Table 2). The tests were broken up into segments by the indications used to order the test. Out of these total costs, $24,180 to $40,920, or approximately 42%, were from testing inconsistent with current expert-based guidance. Cost per positive tests were also identified in each segment. The relationship between each testing segment’s low-cost per positive test estimate and share of testing was represented in a bubble graph (Figure 3). The size of each bubble was proportional to a segment’s positive result rate. The lowest cost per positive test ($758.33 to $1,283.33) was seen when clinician ordering practices were indicated by current expert-based guidance, excluding epidemiologic study data. Though testing consistent with current expert-based guidance, excluding epidemiologic study data, had the lowest cost per positive test; it represented the lowest share of testing (8%). Epidemiologic study had the highest cost per positive test ($7,247.50) and was the indication for half of all testing (50%).

**Figure 3.**
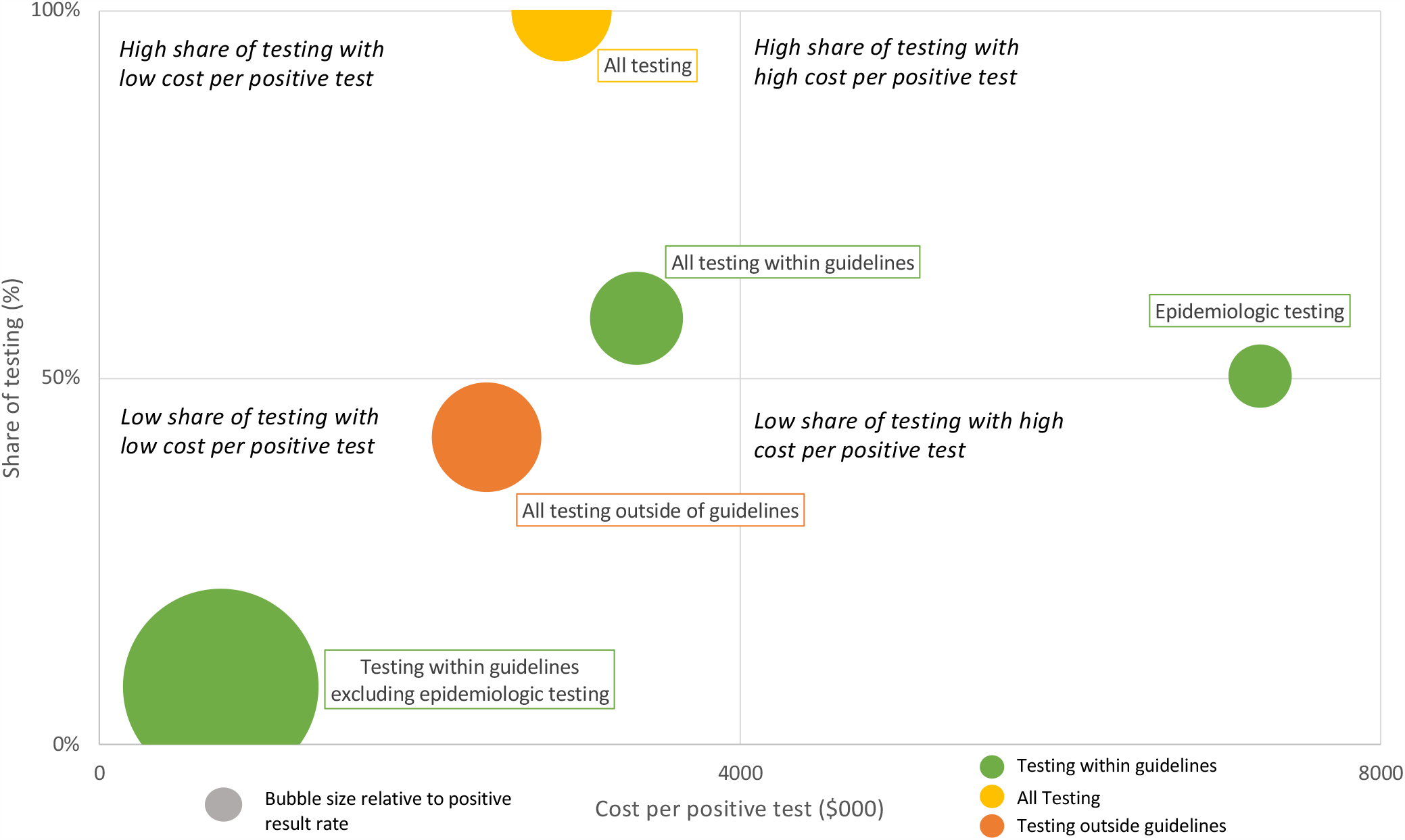
A comparison of the testing scenarios by the cost per positive test and its share of testing. On the x-axis the cost per positive test is represented in US dollars. The share of testing is displayed on the y-axis as a percentage. The relative positive result rate percentage is represented by bubble size.

## Discussion

Apart from public health epidemiology studies of disease prevalence, the most common reason for a clinician to order SARS-CoV-2 antibody testing was to determine presumed immunity in patients with resolved COVID-19 compatible illness. These patients never received SARS-CoV-2 RT-PCR testing at the time of illness. The second most common reason for a clinician to order SARS-CoV-2 antibody testing was to determine presumed immunity in asymptomatic patients believed to have had a past exposure or contact with a person with COVID-19 compatible illness. Neither of these commonly documented indications are consistent with current expert-based guidance from the IDSA or CDC. Because the rate of positive SARS-CoV-2 antibody testing among those indications inconsistent with current IDSA and CDC guidance was significantly lower than the positive rate among expert-backed indications, such inconsistent testing indications are unlikely to change management decisions or be of much clinical value. The prevalence of these inconsistent testing indications suggests that clinical curiosity and patient preference played a large role in initial testing decisions.

The next most commonly identified indication for SARS-CoV-2 antibody testing was detection of RT-PCR negative cases, especially for patients who presented late in a course of illness and potentially have a viral load below the detection limit of RT-PCR assays or when lower respiratory tract sampling was not possible. This was followed by testing to help establish a diagnosis when patients presented with late complications of COVID-19 illness and testing to identify convalescent plasma donors. All three are indications consistent with expert-based guidance from the IDSA or CDC. This was followed by an indication of supporting a diagnosis of COVID-19 in a currently symptomatic patient in the absence of RT-PCR testing and to determine immune status (e.g. return to work, school use) outside of epidemiologic investigation. Both of these indications are inconsistent with current expert-based guidance from the IDSA or CDC.

Two patients with prior positive SARS-CoV-2 RT-PCR tests received antibody testing during prolonged inpatient stays of 18 and 25 days. After initial clinical improvement, both patients had intermittent fever without a clear source. SARS-CoV-2 antibody testing, positive in both cases, was obtained in order to provide supporting evidence of a prior history of COVID-19 infection rather than active SARS-CoV-2 infection or reinfection as a source of the new fevers. This reason for testing is inconsistent with the indications described in this study; whether it represent a valid and clinically meaningful use of the SARS-CoV-2 antibody test remains to be determined.

Finally, we assessed total cost estimates for each testing indication and cost per positive test. “Runaway costs” for the COVID-19 pandemic have been a major concern for the global community – with laboratory testing a key factor.(26–28) In this study, clinical testing consistent with current expert-based guidance had the lowest cost per positive test. Yet, almost half of the total costs were due to testing indications inconsistent with current expert-based guidance. If testing were to continue at the same rate for one year, these inconsistent testing indications would account for almost $500,000 in our hospital. Fortunately, stewardship of laboratory testing is not a new concept.(18,29) Well-designed and carefully thought-out testing strategies have been shown to help increase the awareness of limitations (i.e., false positivity and negativity rate) and appropriateness of a specific test. A possible evidence-based approach to control unnecessary SARS-CoV-2 antibody testing may be in the development and implementation of clinical decision support (CDS) tools that leverage the EMR.(30) This would restrict clinicians to ordering the test under options of CDC and IDSA guidance only until more evidence becomes available.

This study has some limitations. First, retrospective review of clinical documentation is inherently subjective. To counter this limitation, we maintained a conservative threshold for including any single testing indication within a given category. In this study, 11% of testing indications were categorized as “no indication provided,” a frequency consistent with other published literature reliant on retrospective chart review.(31) Second, the duration of antibody response following infection is unclear and asymptomatic individuals may not develop a positive antibody response.(14) Third, IDSA and CDC guidance on testing may change based on evolving literature. Finally, there are concerns about increased false-positive antibody tests due to the low prevalence of COVID-19 in our patient population. However, the use of the EUA Abbott Architect SARS-CoV-2 IgG antibody test in this study provides stronger assurance than previously describe studies.(32,33)

In summary, this investigation identified clinician-documented indications for SARS-CoV-2 antibody testing immediately following testing availability. To our knowledge, we are the first to describe these testing indications from the standpoint of the ordering clinician during the COVID-19 pandemic. Our findings demonstrate a dissociation between clinician described indications for testing and expert-based guidance and a significantly different rate of positive testing between these two groups. Clinical curiosity and patient request appear to have played a significant role in testing decisions. Perhaps this should be expected given the lack of established clinical testing guidelines and the inherent fear and uncertainty driven by the global COVID-19 pandemic. However, without evidence-based guidance, further expansion of SARS-CoV-2 antibody testing availability for clinical purposes may lead to higher healthcare costs with unclear benefit.

## Data Availability

N/A

